# Etiological Profile and Resistance of Bacterial Isolates from Samples of Cerebrospinal Fluid in Kosovo, during the years 2014-2019

**DOI:** 10.1101/2020.08.10.20171900

**Authors:** Rita Qarkaxhija

## Abstract

Cerebrospinal fluid is a dynamic, metabolically active substance that has numerous important functions. It is considered a valuable diagnostic aid in assessing infectious inflammatory conditions, involving the brain, spinal cord, and meninges.

The discovery and use of antimicrobial agents has fundamentally changed medicine from a therapeutic point of view, enabling the treatment of many diseases once considered threatening and deadly. However, after prolonged exposure of various microorganisms to these agents, the development of antimicrobial resistance has been enabled through the adaptive selection mechanism. The emergence of this resistance in the main pathogenic microorganisms is a very problematic and threatening issue in public health, making it a global problem. Resistance of various microorganisms in Kosovo, according to annual reports of CAESAR, compared to other European countries, is quite worrying. It reaches the tops of the lists for high resistance, along with other Balkan countries, such as Serbia, Montenegro, and North Macedonia.

The purpose of this research was to define the etiology and level of resistance of microorganisms to antimicrobial agents, which are encountered in invasive samples of cerebrospinal fluid. This research is a retrospective, descriptive type analysis, which includes the data gathered from January 1, 2014 to May 7, 2019. We have a total of 185 bacteria isolated from 1499 isolates, conducted at the National Institute of Public Health in Kosovo. The determination of antimicrobial resistance was performed according to automated systems and the method of disk diffusion.

## 1. Introduction

### 1.1. Cerebrospinal Fluid, Production, Resorption, and Diagnostic Importance

Cerebrospinal fluid is a colorless, odorless fluid that travels through the ventricular spaces and into the subarachnoid space. Studies of cerebrospinal fluid (CSF) began in the 1800s, when Quincke introduced and developed the lumbar puncture method. The cerebral ventricles are the largest intracranial spaces, containing the cerebrospinal fluid. The choroid plexus is the main site of production of cerebrospinal fluid. The choroid plexus is a network of blood capillaries and ependymal cells, located in the ventricular system [1]. In the adult brain, the choroid plexus is a well-vascularized papillary structure. Meninges are the three membranes that completely surround the central nervous system (i.e the brain and spinal cord). Their main function is to protect the central nervous system. These three membranous layers are: Dura mater, Arachnoid mater, Pia mater.

Cerebrospinal fluid is not simply a plasma ultrafiltrate [2]. Its production and secretion are highly specialized processes, performed mainly by the choroid plexus. The hypothesis for this fact was first raised by the neurosurgeon Walter Dandy, who as a treatment for hydrocephalus used the removal of the choroid plexus [3]. Less than 10% of CSF is derived from extra choroidal sources. The total volume of cerebrospinal fluid in adults has a range of 140 ml to 270 ml, where 25% fills the ventricles, and the rest circulates through the basal cisterns, subarachnoid space, and around the spinal cord. About 600 ml of CSF are formed during the day, which means that this fluid is replaced 4 times a day within 24 hours [4]. The first step in the production of CSF from the choroid plexus, consists in the passive filtration of plasma from the choroidal capillaries, according to a pressure gradient. The second step is based on active transport, from the interstitial compartment to the ventricular lumen, along the choroidal epithelium.The CSF content is slightly acidic, which in turn, reflects the increased partial pressure of carbon dioxide (pCO2), and suggests a reduced buffering capacity of CSF compared to plasma. Normally, in CSF there are no more than 5 cells per millimeter. Also, in the physiological state the levels of amino acids and glucose in CSF are lower than in plasma. Recent studies, using the latest methods of sensitive protein identification, have identified over 2500 unique proteins, of which only 400 have been found in the ‘human plasma protein’ database [5]. Any increase in intraventricular pressure (whether acute or chronic) lowers the pressure gradient along the hematoencephalic barrier, and this lowers plasma filtration. This shows that the secretion of CSF is regulated in such a way that it leaves no room for errors.Circulation of CSF is a dynamic phenomenon, which plays a key role in the homeostasis of cerebrospinal fluid. The flow of CSF is pulsatile, corresponding to the systolic pulse of the choroidal arteries. The fluid flows in a rostrocaudal direction.In contrast to the secretion of CSF, which is a highly orchestrated process, the resorption of CSF occurs through the physiology of “bulk flow” (one of the three processes of exchange in capillaries, through which proteins move through various cellular components), a process of which is not regulated. The resorption process occurs in the sagittal cranial sinus mainly, but a small part of the resorption is also carried out by the choroid plexus.

Cerebrospinal fluid, being a metabolically active and dynamic substance, has a valuable and irreplaceable role in diagnostics.It helps evaluate inflammatory, infectious, and non-infectious conditions, including the brain, spinal cord, and meninges. In addition to helping in assessment of inflammatory conditions, CSF analysis helps us diagnose autoimmune diseases, CT-negative subarachnoid hemorrhage, and detect leptomeningeal metastases [6]. CSF sampling is done by lumbar puncture [7].

### 1.2. Infectious Diseases Associated with the Presence of Bacteria in CSF - Bacterial Meningitis

In bacterial infections of the central nervous system, the conclusions drawn from CSF are considered to be essential in confirming a diagnosis, identifying the causative microorganisms, and guiding antimicrobial therapy. In the CNS, bacterial infections can cause: Bacterial meningitis, brain abscess, encephalitis, epidural spinal abscess. But, only bacterial meningitis will be discussed here, as it is one of the most common and dangerous infections, where the earlier the diagnosis is made, the lower the mortality.

Meningitis is called inflammation of the meninges, subcutaneous structures and CSF. It is one of the most dangerous infections [8]. Based on the onset of symptomatology, bacterial meningitis is classified into:

1. Acute meningitis (< 24 hours)
2. Subacute meningitis (1-7 days)
3. Chronic meningitis (> 7 days)

Bacterial meningitis has no common epidemiological features. The vast majority of them are in fact sporadic [9]. Only meningococcal meningitis and meningitis from Haemophilus influenzae, which have a sporadic, endemic and epidemic character. Epidemics of meningitis from Escherichia Coli have also been encountered, but they are mostly nosocomial. According to the Wold Health Organisation (WHO), in 2010, bacterial meningitis occurred in 3 out of 100,000 inhabitants, as opposed to viral meningitis which was encountered in 11 inhabitants out of 100,000 inhabitants [9].

Morbidity and mortality depend on the cause, the age of the patient, as well as the health condition of the patient [10]. The etiological causes of bacterial meningitis are numerous. These etiological causes, according to the place of contraction of bacteria are divided into: Etiological causes acquired in the community, and Nosocomial etiological agents (acquired in hospital settings). Whereas, depending on the degree of resistance of the respective microorganisms are divided into: Bacteria resistant to many drugs, and non-drug resistant bacteria.

The WHO in 2017 released a list of the 12 most dangerous bacteria of recent years, which are gaining resistance to almost all types of antimicrobials we possess. Unfortunately, of those 12, there are 7 that can cause bacterial meningitis and septicemia [11]. Bacterial meningitis is a disease that is usually acquired hematogenously. The pathways through which bacteremia occurs are multiple, and those bacteria sometimes take the path to the subarachnoid space. From the blood, bacteria enter the large venous sinuses in the brain. In these venous canals, the blood moves slowly, so bacteria can settle easily. Bacteria then penetrate the dura, the arachnoid, and thus infect the CSF [8]. Rarely, bacteria penetrate through a defect in the cribriform plate of the ethmoid bone, or even through a small opening, which usually remains behind fractures of the basal part of the skull. Patients who develop otitis or even a sinus infection, easily develop brain abscess, and from the abscess to the brain, the infection can easily spread per continuitatem to the subarachnoid space. As the blood-brain barrier blocks the passage of immunoglobulins and complement, bacteria develop smoothly and multiply rapidly at first (as they are not detected by the host’s defense system) [10]. As the bacteria multiply, polymorphonuclear cells (neutrophils and macrophages) go to the scene. Then, while trying to defeat microorganisms, release enzymes, cytokines and toxic oxygen products [8]. In this way the cerebral microvasculature is damaged, which leads to the breakdown of the blood-brain barrier. The characteristic triad of clinical presentation of meningitis is: Elevated temperature, headache, neck rigidity [10].

Other common clinical symptoms include mood swings, nausea, vomiting, and photophobia [10].Diagnosis is based on clinical and microbiological criteria (quantitative culture). In choosing the appropriate antimicrobial drug for the treatment of CNS infections, especially meningitis, professional and medical criteria should be considered. In these criteria, in the first place is the identification of the pathogenic microorganism (bacteria in this case), testing of its susceptibility or resistance to antimicrobials, recognition of patient characteristics (which may be the localization of infection, age, immunological and hemodynamic status and the functional state of the organ systems), as well as the economic cost of the antimicrobials [12]. Knowledge of these factors is essential for the selection of the appropriate antimicrobial, because a large number of these microorganisms possess the property of adaptation and development of resistance.

### 1.3. Causes of Antimicrobial Resistance

Antimicrobials are drugs that slow down the growth of microorganisms (whether they are bacteria, fungi, viruses or parasites), or that completely destroy them. Resistance to these drugs occurs when microorganisms adapt to the drug, and continue to replicate in its presence. This has catastrophic consequences. Antimicrobial resistance (AMR) is one of the most dangerous global health threats. In recent years, antimicrobial resistance has been compared to the risk of global warming because both are international problems that do not recognize state borders. It is estimated that every year, worldwide, 700,000 people die from infections resistant to therapy, and by 2050 this number will reach 10 million [13].The source of the problem, and the effective approach to it involves many state authorities, services and public service institutions. The solution to the problem affects many sectors, from human to veterinary health, and also depends on agriculture and the environment in general.Essential to address antimicrobial resistance (especially antibacterial) effectively are: supervision, monitoring, prevention and control of antibiotics, awareness, education and financial stability [14]. Antimicrobials, and especially antibiotics, have not only saved the lives of millions of people, but have also helped in the development of medicine and surgery. The causes of antibiotic resistance are:

#### 1.3.1. Overuse of antibiotics

Despite the alarm to prevent the overuse of antibiotics, they are being used recklessly.Antibiotics are removing drug-sensitive bacteria from the race, leaving only those resistant bacteria to predominate, as a result of natural selection.In many countries, such as the US, taking antibiotics is more regulated and monitored, and as a result they cannot be taken without a doctor’s prescription. While in many other countries, antibiotics can be bought and used by anyone.

#### 1.3.2. Irregular prescription of antibiotics by doctors

Studies have shown that in 30% to 50% of cases one of the following is wrong: the indication for therapy, the choice of therapeutic agent or the duration of treatment [15]. Incorrect description of antibiotics also contributes to the development of resistance. Subinhibitory / subtherapeutic concentrations promote resistance in bacteria by supporting genetic changes in bacterial DNA [16].

#### 1.3.3. Extensive agricultural use

In the developed world and in the developing world, antibiotics are used as growth supplements in livestock. The use of antibiotics in livestock improves animal health, and they will provide more abundant and higher quality products if they are healthy. It is estimated that 80% of antibiotics sold in the US are used for livestock [17]. And the transfer of animal-resistant bacteria to humans was observed more than 35 years ago, when in the intestinal flora of farm animals bacterial-resistant bacteria were observed [17]. When humans consume animal feed, these antibiotics enter their system. The use of antibiotics in livestock, also has a serious effect on the microbiome of the environment, because antibiotics are excreted through urine and feces. In addition to catastrophic applications in livestock, antibiotics are being used continuously in agriculture, as pesticides (tetracyclines and streptomycin) [18].

#### 1.3.4. Lack of new antibiotics

The development of new antibiotics is no longer considered to be a wise investment by pharmaceutical companies. Of the 18 largest companies, 13 have completely abandoned antibiotics [17]. The reasons that pharmaceutical companies no longer invest in antibiotics are: shorter use of antibiotics compared to drugs for chronic diseases, lower price of antibiotics compared to other drugs, storage of newer antibiotics only for cases of resistance multiple, deficiencies in the regulation of patents for antibiotics [19].

#### 1.3.5. Bureaucratic barriers

The FDA (Food and Drug Administration) has made the procedure for approving new antibiotics more difficult. This is because, under the new rules and policies, comparing new antibiotics with placebo is being considered unethical; instead clinical trials are based on proving the lack of inferiority of new antibiotics, compared to old ones [20].

## 2. Purpose of Research

The purpose of this paper, is:

- Research of culture samples taken from the cerebrospinal fluid and determination of their etiological profile.
- Analysis of the results of resistance of bacteria towards antimicrobial agents.
- Critical and comprehensive assessment of the antimicrobial resistance of certain bacterial isolates.
- Comparison of antimicrobial resistance in Kosovo, with countries in the region.
- Comparison of antimicrobial resistance based on bacteria.
- Providing data for drafting a roadmap for preventive measures.
- Establishing the basis for ongoing evaluation in order to improve and meet the criteria of the strategy for resistance to antimicrobial agents.
- Guidance and assistance for the preparation of new directives of the Ministry of Health of Kosovo, for prevention of the negligent use of antimicrobials in hospitals.

## 3. Material and Methodology of Work

The research is a retrospective, descriptive analysis, which includes data from 5 years (from January 1, 2014 - until May 7, 2019). In the National Institute of Public Health (Department of Microbiology),in Prishtina, a total of 1499 cases were tested, where 185 cases were isolated, while in 1314 cases there were no isolates.The research samples are part of the Central Asian and European Surveillance of Antimicrobial Resistance (CAESAR) study, which is a key network of the World Health Organization (WHO), which collects data on antimicrobial resistance testing from samples, for 9 species of bacteria (as of 2018) that are relevant greater in clinic and in public health. At the National Institute of Public Health of Kosovo, antimicrobial resistance is evaluated via automated systems and the method of disk diffusion.All laboratories in Kosovo, since 2013, use the EUCAST method (European committee on antimicrobial susceptibility testing39), as a standard for the analysis and interpretation of antimicrobial susceptibility and resistance testing. These standards are based on criteria set by the European Committee. The results of susceptibility and resistance to antimicrobials are presented as the ratio between the number of cases reported in the specified time period and the values of resistant cases reported as a percentage, for a specific antimicrobial agent, e.g. Total number of isolated E.coli samples, and expressed resistance in percentage to beta-lactam antibiotics (e.g. Aztreonami). Resistance ratios are not rounded, but are presented in decimal places. The processing of statistical analysis for the data collected, was done using the Python programming language and its Matplotlib and Numpy libraries. As the study is for a period of 5 years, EUCAST guidelines have changed (e.g. new antibiotics have been added for testing, or some antibiotics have been removed), resulting in slight discrepancies in the data. But we can not rule out the possibility of technical errors (human error).

## 4. Results

A total of 1499 cases were tested, during the period 1 January 2014 to 7 May 2019, of which in 185 samples relevant microorganisms were isolated, while in 1314 samples microorganisms were not isolated. The data are presented below, in tabular and graphical form. It should be noted that the percentages presented in the tables reflect the number of tests, and not necessarily the real percentage of resistance in the population.

From the data in Table 1, it can be concluded that the most common cases of microorganisms isolated from CSF are those of Acetinobacter spp (59 cases in total, or 31.89 percent, of which Acetinobacter baumannii, with 44 cases, or 74.57 percent). Immediately in second place are cases of Staphylococcus spp., where 26 cases have been isolated, or 14.05 percent, where the most common of them was Staph. Aureus, with 2 cases (or 7.6 percent). In the third place are samples where Streptococcus species were isolated (23 cases, or 12.43 percent), and the most common of them was Strep.Pneumoniae- with 9 cases (or 39.13 percent). Pseudomonas spp rank on the fourth place, with 15 of them (or 8.1 percent), of which P.aeruginosa stands out with 12 cases (or 80 percent).

**Table 1:**
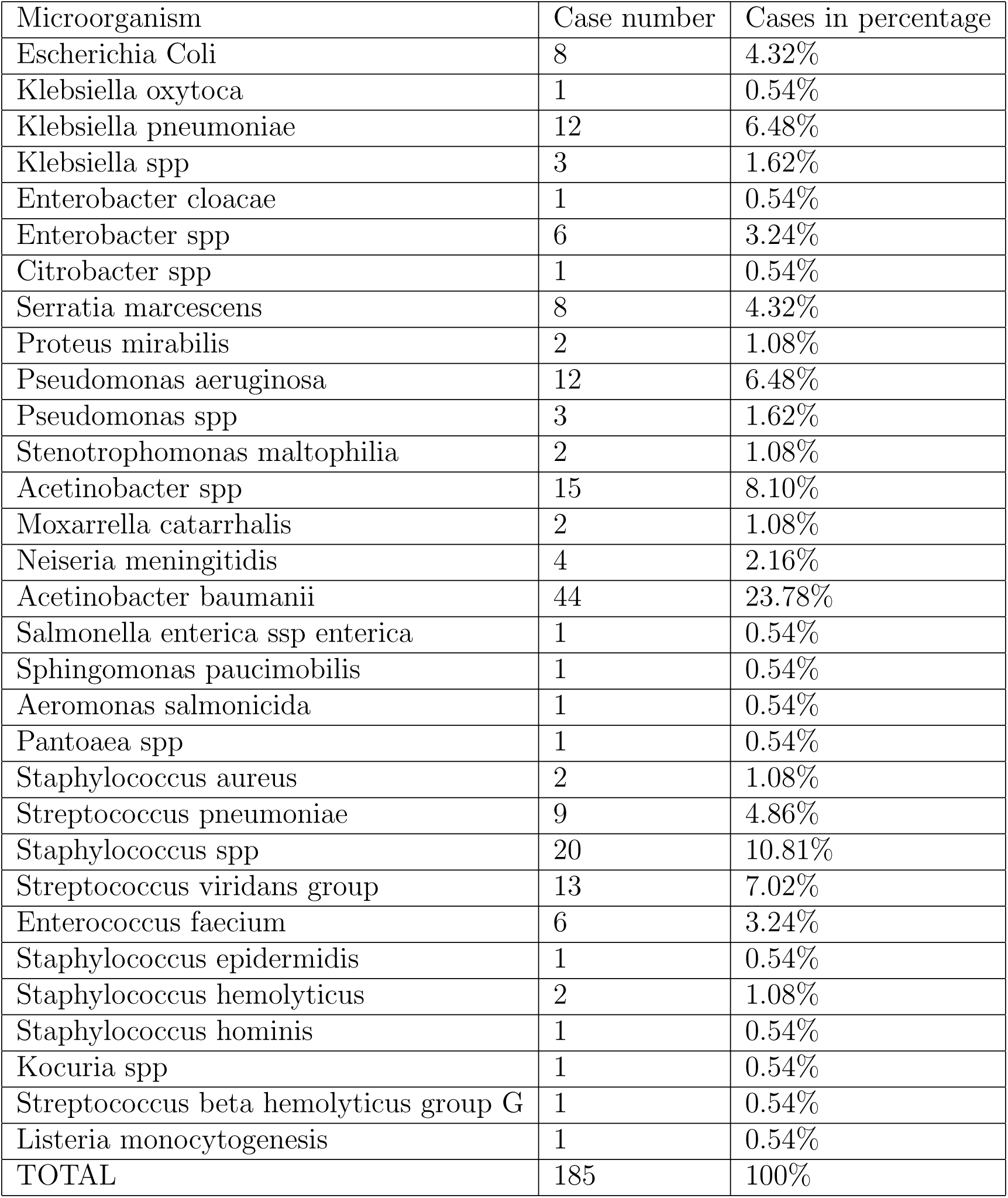
Etiological profile presented in numerical value and in percentage, in relation to the total number of isolated samples.

**Figure 1:**
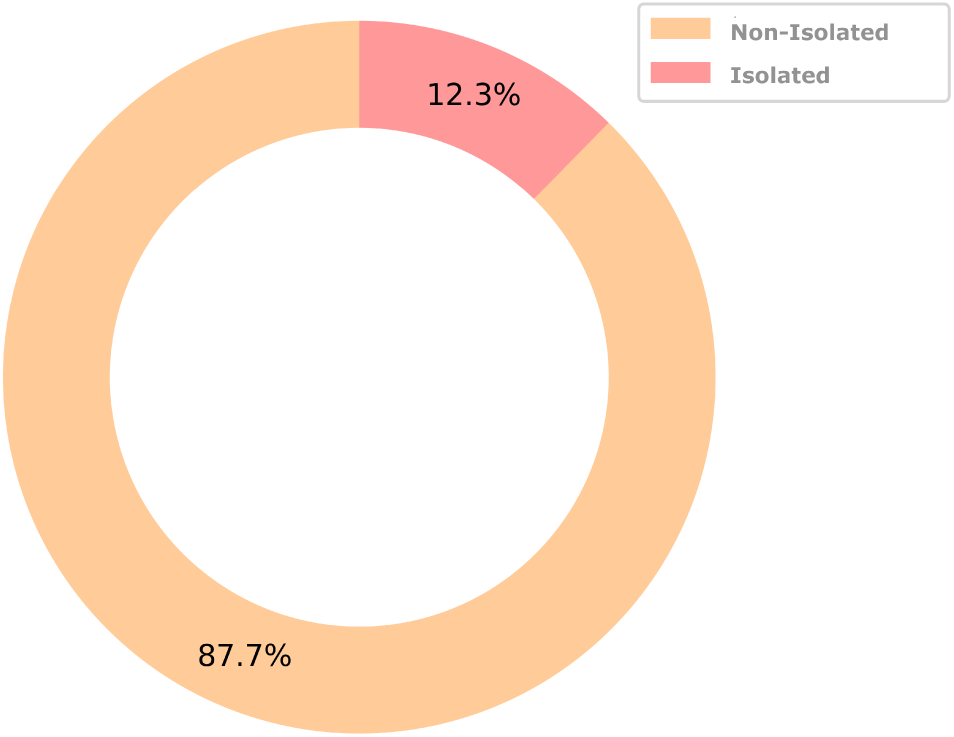
Schematic presentation of results

Looking at the figure 3, we see that E.Coli is highly resistant to the Aminoglycoside class of antibiotics (62.5% in Gentamicin, 62.5% in Tobramycin). Significant resistance is also seen in third generation cephalosporins (62.5% in Ceftriaxone, 62.5% in Ceftazidime and 62.5% in Cefotaxime). of 8 in total (62.5%).

**Figure 2:**
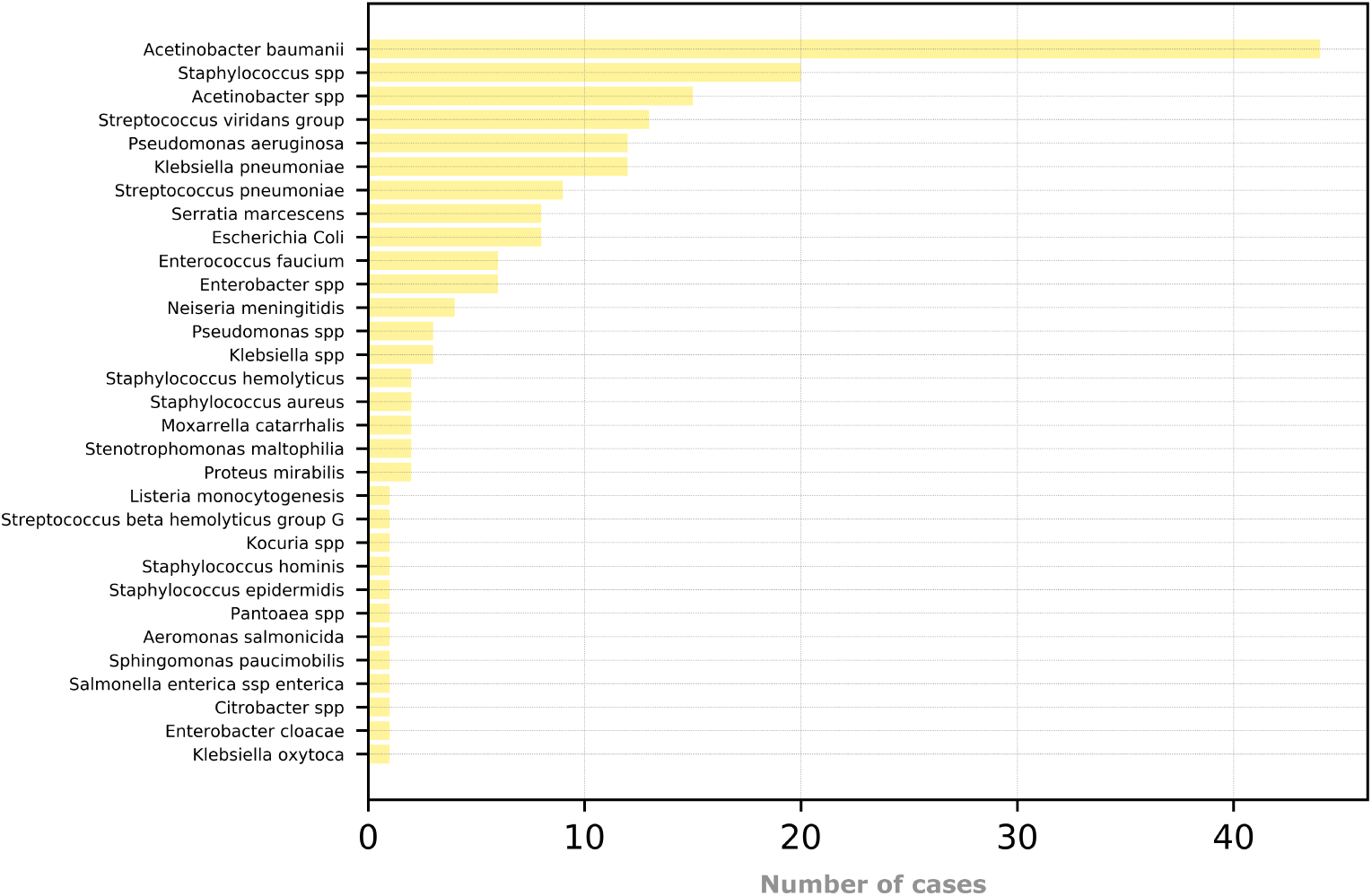
Schematic presentation of the total number of bacterial isolates, obtained from National Institute of Public Health, for the period 2014-2019

**Figure 3:**
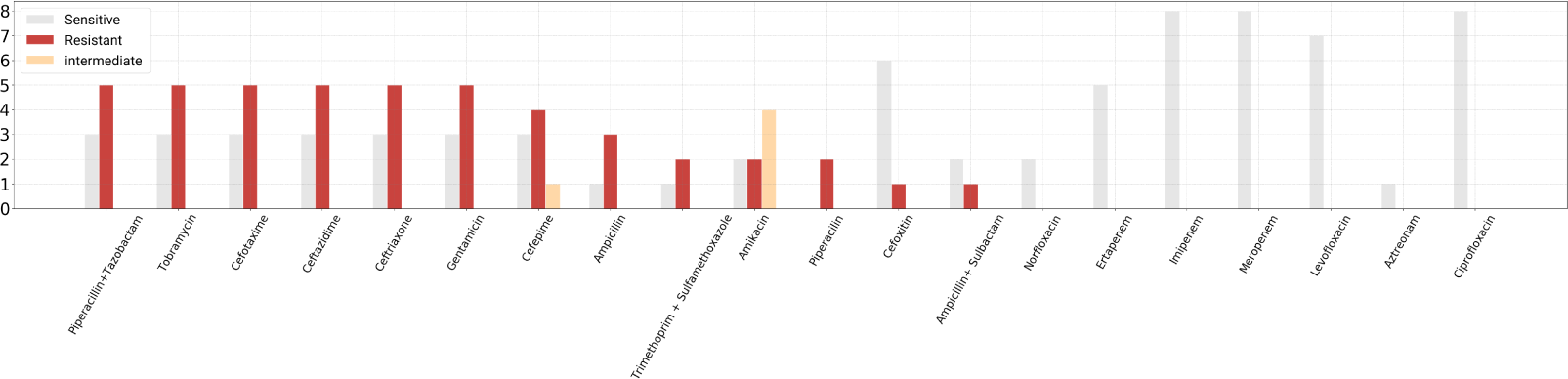
Levels of resistance and sensitivity of E.Coli to relevant antimicrobial agents

According to the graphical presentation in the figure 4, K.Pneumoniae shows resistance mainly to the third generation Cephalosporins (Cefotaxime with 83.3%, Ceftriaxone 81.82%, Ceftazidime 50%). Also, resistance is seen in the antimicrobial combinations: Ampicillin + Sulbactam and Trimethoprim + Sulfamethoxazole, where resistance is seen with 83.3% and 100% respectively.

**Figure 4:**
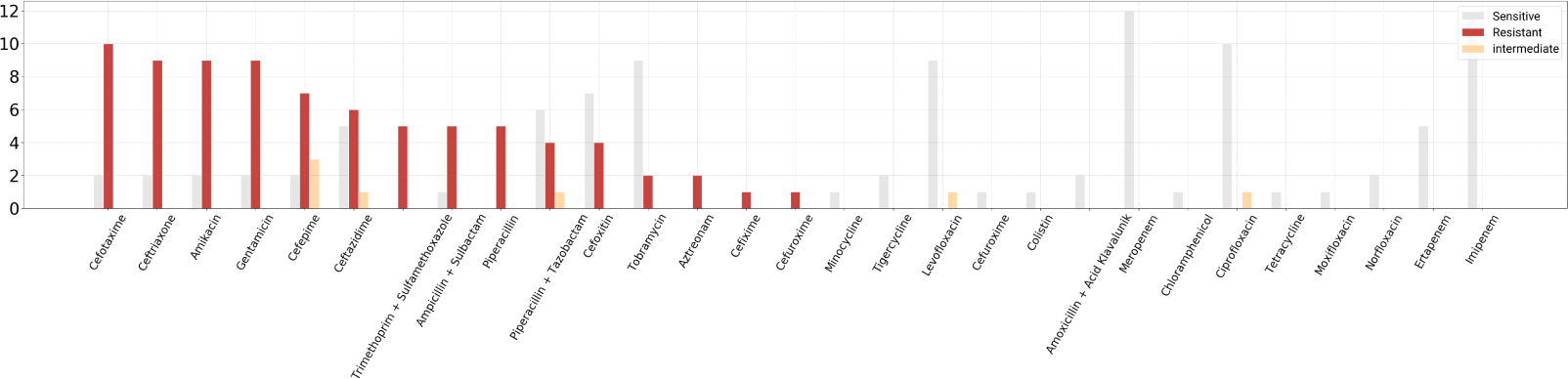
Levels of resistance and sensitivity of K.pneumoniae to relevant agents

Klebsiella spp, as shown on figure 5 were seen to be resistant to third generation Cephalosporins (Cieftriaxone with 66.67%, Cefotaxime with 66.67% and Ceftazidime with 66.67%). The same resistance number is also observed in the semi-synthetic aminoglycoside antibiotic, Amikacin.

In Enterobacter spp.as seen on figure 6, high levels of resistance to third-generation Cephalosporins were seen (Ceftriaxone with 83.33% and Cefotaxime with 83.33%). The same resistance number (83.33%) is also encountered against the aminoglycoside Gentamicin, and against the broad spectrum beta-lactam antibiotic, Piperacillin.

**Figure 5:**
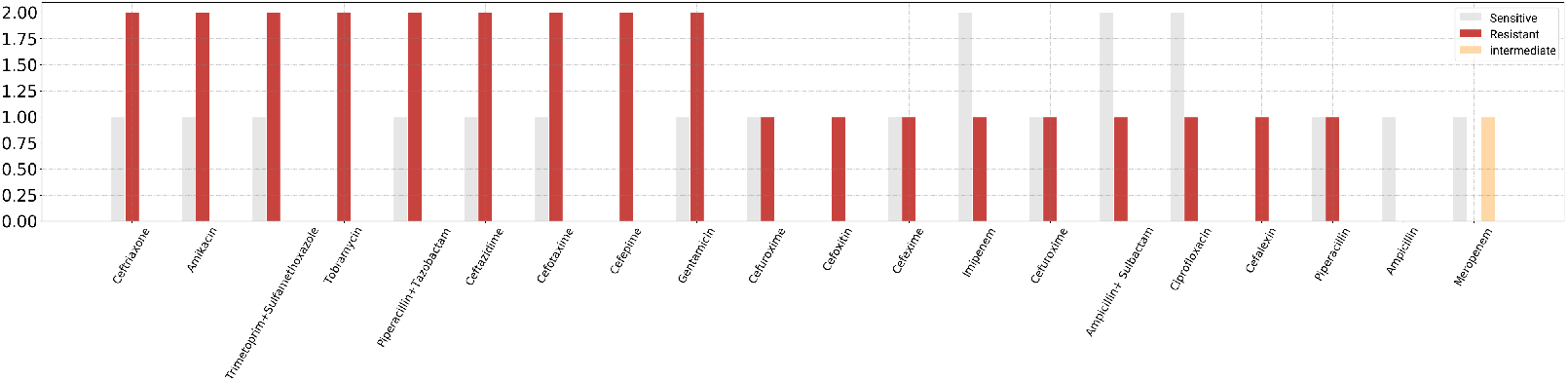
Levels of resistance and sensitivity of Klebsiella spp to relevant agents

**Figure 6:**
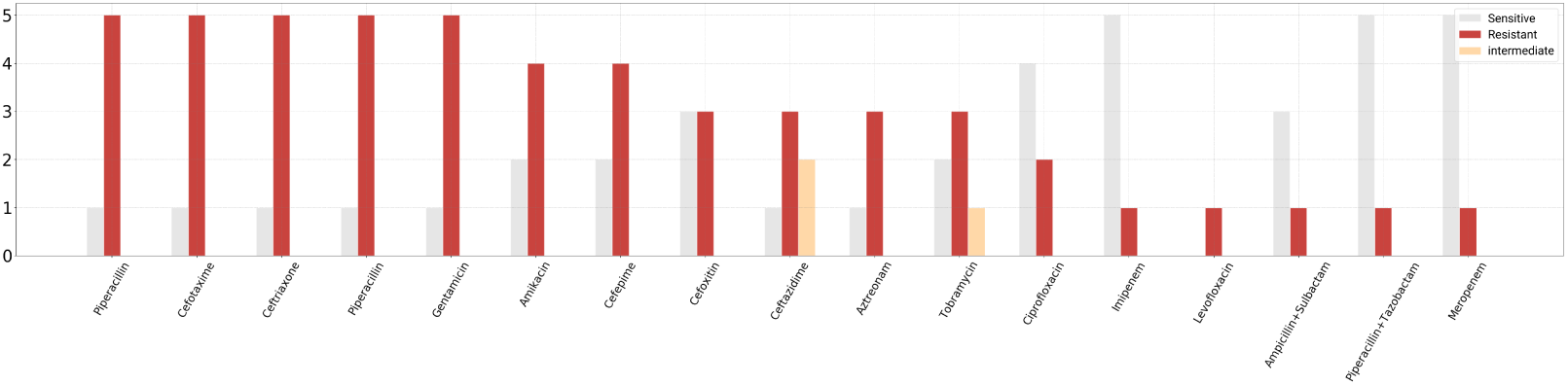
Levels of resistance and sensitivity of Enterobacter spp to relevant agents

Serratia marcescens, as shown on figure 7, being an opportunistic microbial pathogen, has been at the center of nosocomial infections in recent years [21]. The resistance, found in the 8 isolated samples, especially in the second and third generation Cephalosporins, is worrying.

**Figure 7:**
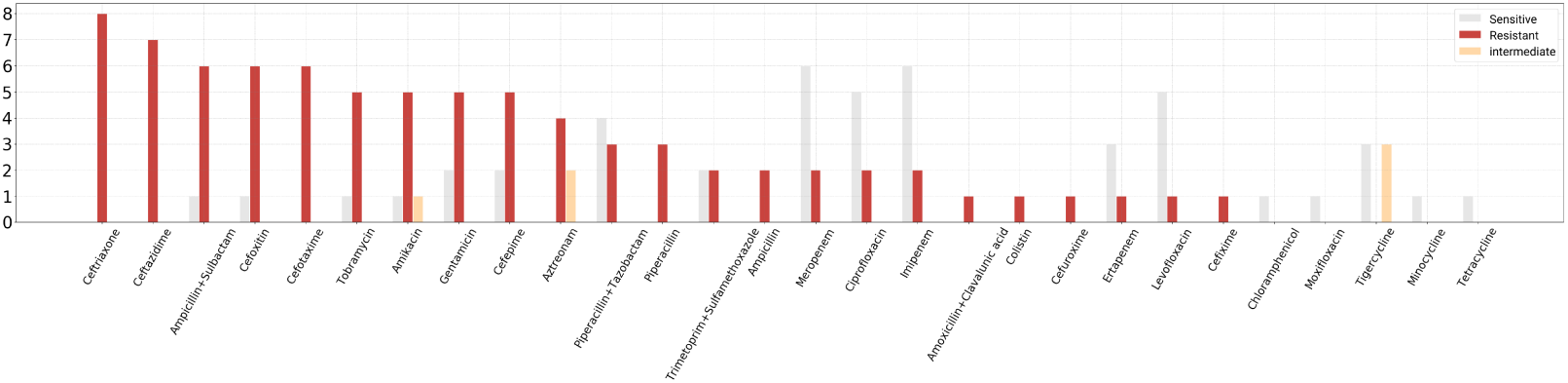
Levels of resistance and sensitivity of Serratia marcescens to relevant agents

In samples where Proteus mirabilis was isolated, the highest resistance was found towards Cephalosporins (Cefoxitin, Ceftriaxone, Cefotaxime and Ceftazidime) as well as towards the TMP / SMX combination (Figure 8).

**Figure 8:**
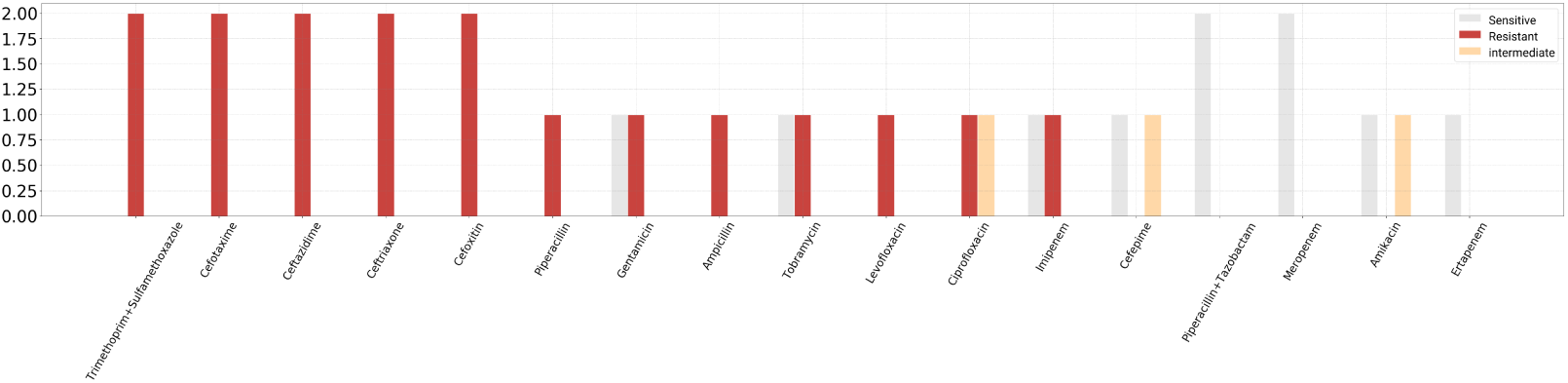
Levels of resistance and sensitivity of Proteus mirabilis to relevant agents

In a total of 12 isolated cases of Pseudomonas aeruginosa, a marked resistance to the carbapenem antibiotics was encountered (Imipenem with 83.33% and Meropenem with 83.33%), as well as towards cephalosporin (second and third generation) antibiotics, as shown on figure 9.

**Figure 9:**
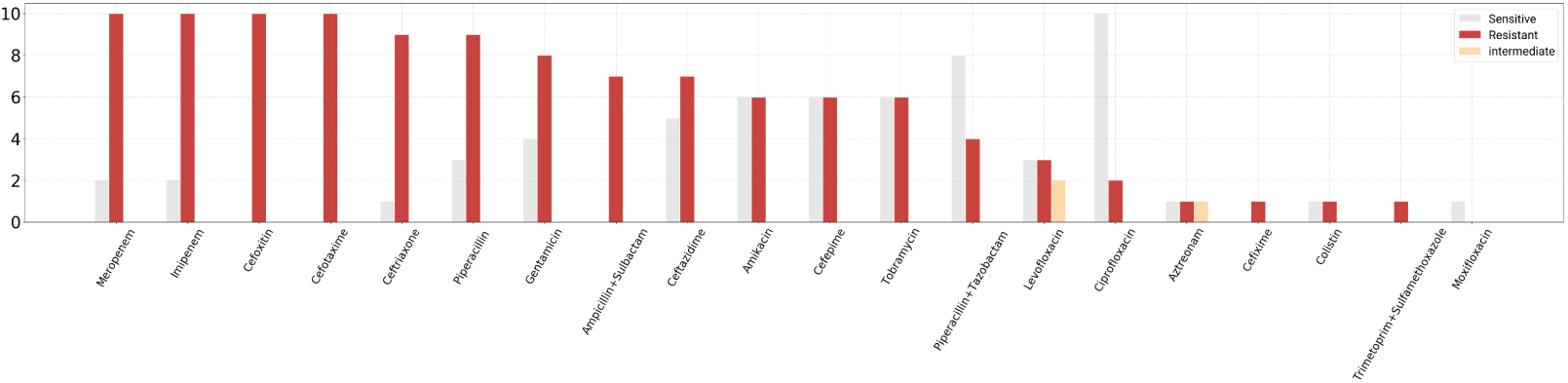
Levels of resistance and sensitivity of P.aeruginosa to relevant agents

As shown on figure 10,resistance of Pseudomonas spp. was mainly en-countered in Aminoglycosides and Cephalosporins, unlike P.aeruginosa where resistance was mainly encountered in Carbapenems.

**Figure 10:**
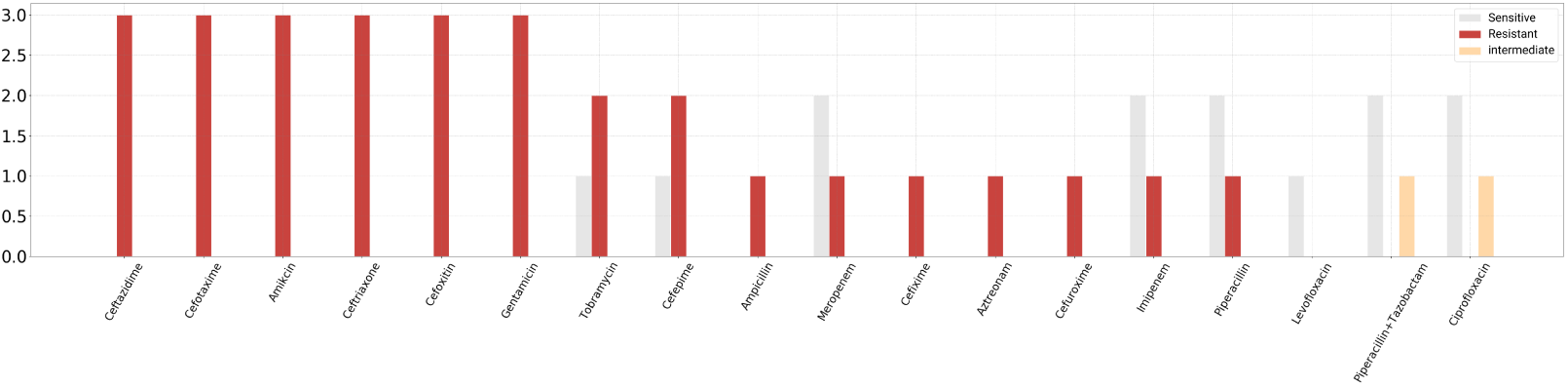
Levels of resistance and sensitivity of Pseudomonas spp to relevant agents

The unusual bacterium, Stenotrophomonas maltophilia, seems to have stayed true to its reputation as “multidrug-resistant” this time around, proving very difficult to treat. Most strains of Stenotrophomonas maltophilia are characterized by their resistance to many broad-spectrum antibiotics, including the class of Carbapenems [22], as was the case in these two samples isolated from cerebrospinal fluid. As shown on figure 11,we also encounter marked resistance towards Aminoglycosides as well as Cephalosporins.

**Figure 11:**
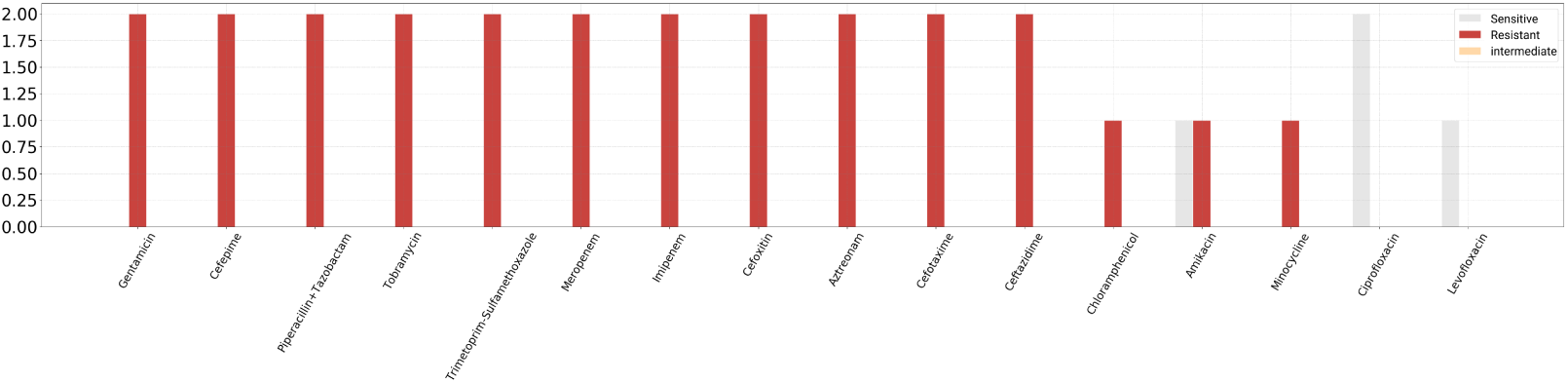
Levels of resistance and sensitivity of Stenotrophomonas maltophilia to relevant agents

Acetinobacter spp being nosocomial pathogens, demonstrate high numbers of resistance. Out of a total of 15 isolated cases in CSF samples for the last 5 years, resistance (as seen on figure 12) was encountered towards Aminoglycosides and Carbapenems. The Cephalosporins were not left far behind either, although the resistance towards them was slightly smaller than in the other two classes of antibiotics mentioned above.

**Figure 12:**
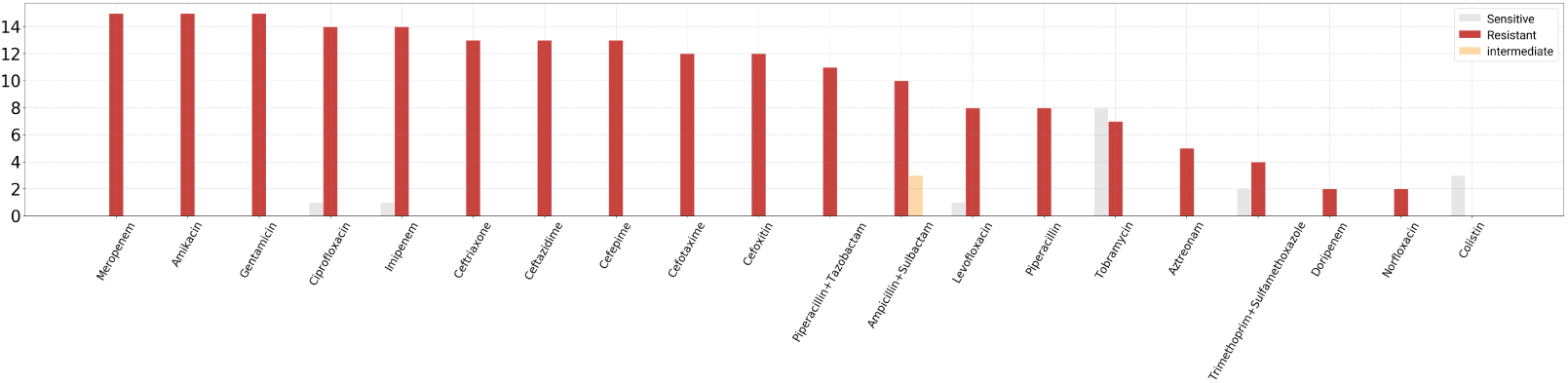
Levels of resistance and sensitivity of Acetinobacter spp to relevant agents

The gram-negative diplococcus, Moraxella Catarrhalis, was encountered only 2 times over a period of 5 years, and the highest level of resistance (as seen on figure 13) was encountered towards Amoxicillin - the broad-spectrum semisynthetic antibiotic, an ampicillin analogue.

**Figure 13:**
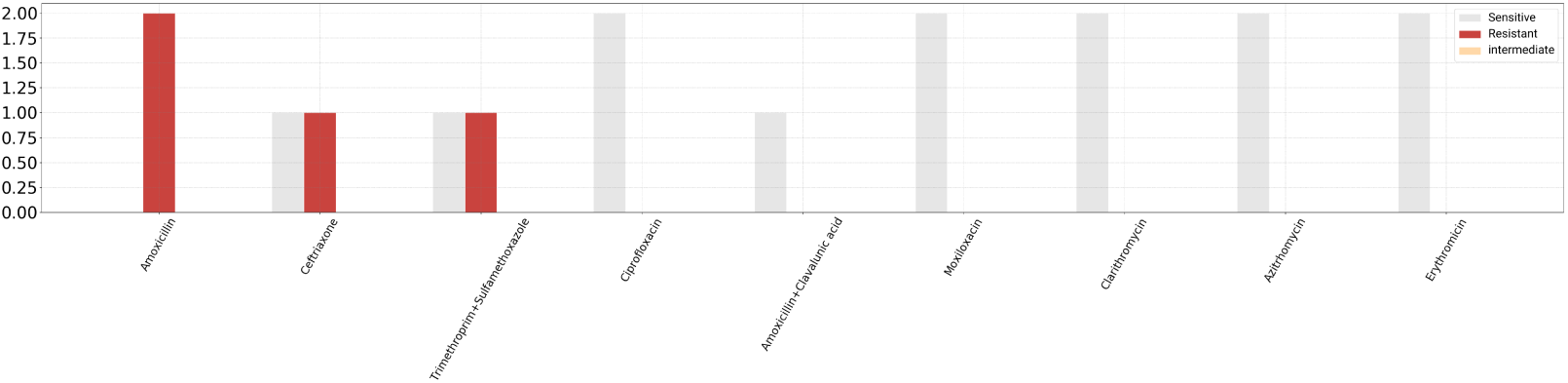
Levels of resistance and sensitivity of Moraxella catarrhalis spp to relevant agents

No major resistance of Neiseria meningitidis towards antibiotics has been encountered. Of our 4 cases (figure 14), resistance was encountered only towards Kolistin, which for the last 50 years has been considered as “last line therapy” [23].

**Figure 14:**
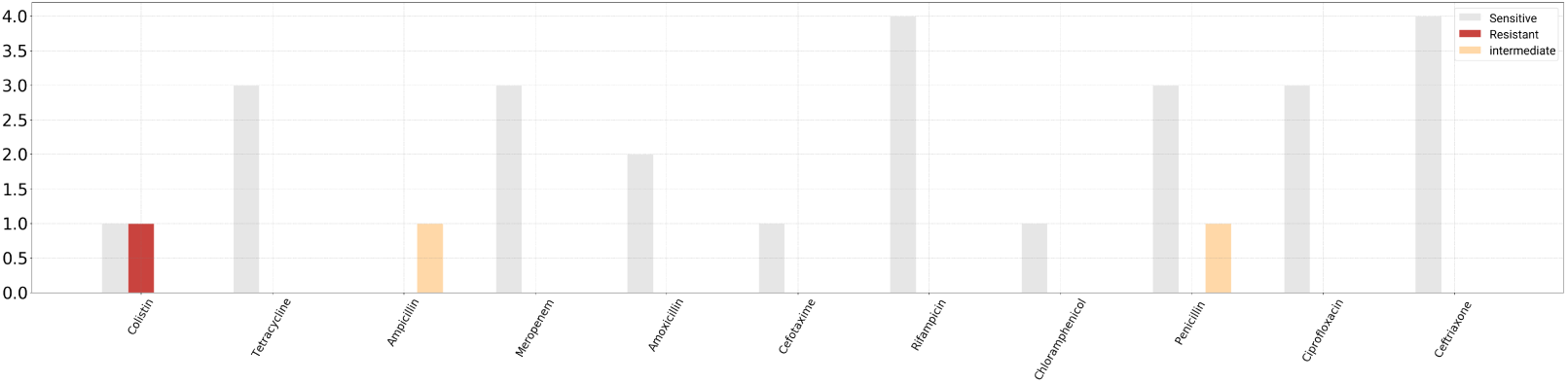
Levels of resistance and sensitivity of N.Meningitidis spp to relevant antimicrobial agents

Acetinobacter baumanii, without a doubt, was the most isolated bacterium during this 5-year period from CSF samples (44 such cases in total). There are two reasons for this: it is a pathogenic opportunistic bacterium, and it is encountered in hospital environments (nosocomial infection) 43. The most pronounced resistance, as seen on figure 15 was found towards Carbapenems (Imipenem with 97.73%, Ertapenem with 100%, Aztreonam with 100%), Cephalosporins (Cefixime, Cefuroxime, Cefoxitin, Ceftazidime, Ceftriaxone - all with 100%), Aminoglycosides with 100%, Tobramycin (100%), Quinolone (Moxifloxacin 66.67%, Norfloxacin 100%) and Fluoroquinolone (Levofloxacin 100%).

**Figure 15:**
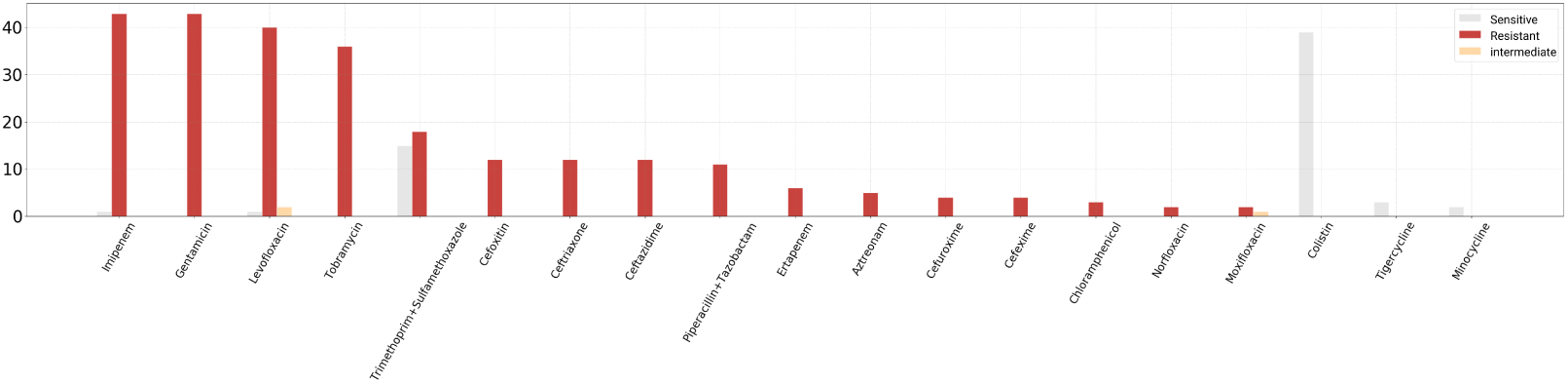
Levels of resistance and sensitivity of Acinetobacter baumannii spp to relevant agents

In total,two cases were encountered, where Staphylococcus aureus was isolated. This bacterium causes 0.3% −8.8% [24] of infections worldwide, and is estimated to be the second most common cause of CSF shunt infections, being surpassed only by Staphylococcus epidermidis [24]. In our 2 samples with Staphylococcus aureus,as shown on figure 16 it was found to be highly resistant to the macrolide group of antibiotics (100% resistance to Clarithromycin, Azithromycin and Erythromycin). Also, resistance is encountered in the antibiotic Penicillin (100% of cases).

**Figure 16:**
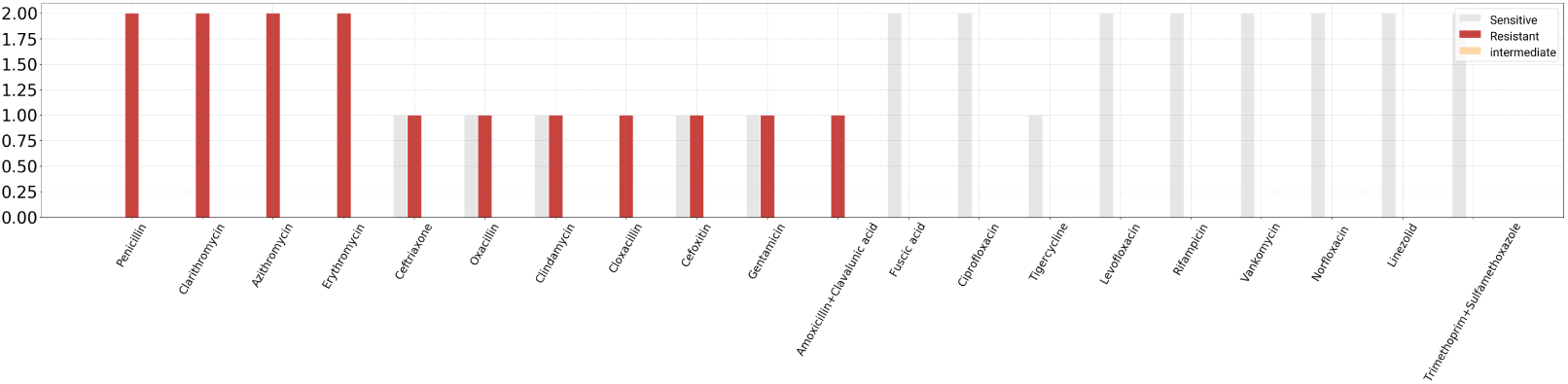
Levels of resistance and sensitivity of Staph.aureus to relevant agents

Out of a total of 9 isolated cases of Streptococcus pneumoniae, the highest resistance (as shown on figure 17) was encountered in the TMP + SMX combination (50% of cases turned out to be resistant). Also in 33.33% of cases we had resistance to the antibiotics Clindamycin, Erythromycin, Cloxacillin and Norfloxacin.

**Figure 17:**
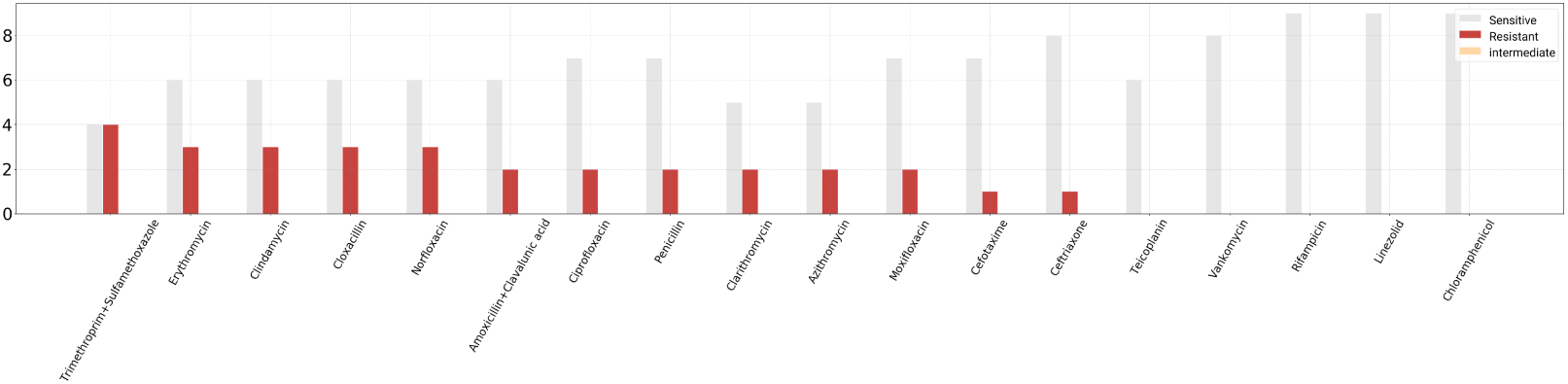
Levels of resistance and sensitivity of Streptococcus pneumoniae to relevant agents

In total, during this period, 20 cases were collected where Staph. Spp was isolated from CSF. Of these cases the highest resistance,as seen on figure 18, (95%) was encountered towards Penicillin. Macrolide antibiotics Clarithromycin and Azithromycin with 77.78% resistance, were not left far behind.

**Figure 18:**
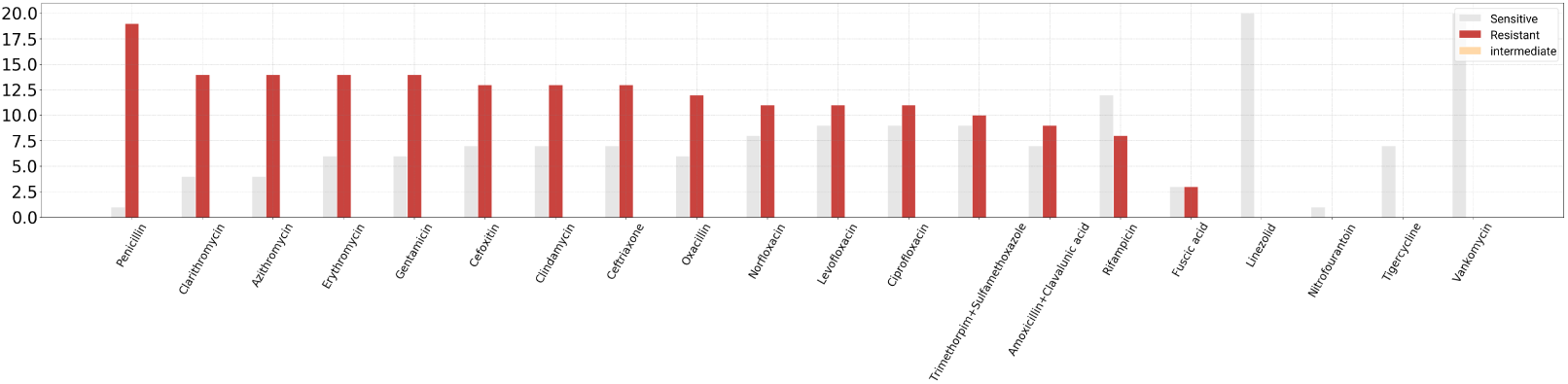
Levels of resistance and sensitivity of Staphylococcus spp to relevant agents

Streptococcus viridans, is the normal flora of the human mouth [25]. Although their pathogenicity is generally low [26] during this 5-year period, a total of 13 cases of streptococcus viridans have been encountered in patients(Figure 19). The most prominent resistance was encountered in the TMP + SMX combination, in 90.91% of cases. Subsequently resistance towards Penicillin in 46.15% of cases was encountered, as well as in Ampicillin in 55.56

**Figure 19:**
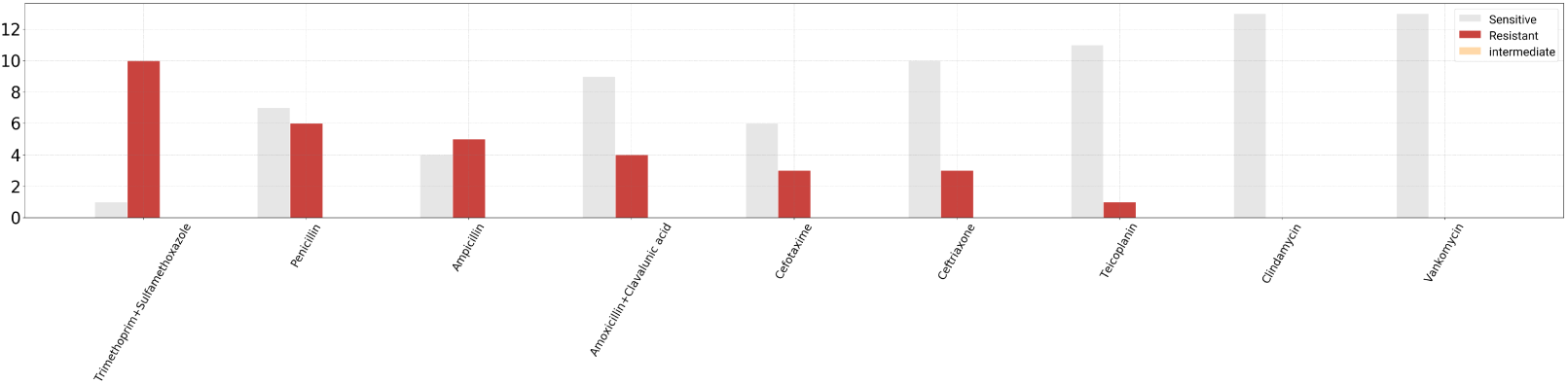
Levels of resistance and sensitivity of the group of Streptococcus viridans to the respective agents

This gram-positive, hemolytic alpha bacterium-Enterococcus faecium, is considered to be one of the most dangerous bacteria for two reasons: it causes neonatal meningitis, neonatal sepsis and is constantly gaining strong resistance to vancomycin [27]. In 6 cases encountered in the University Clinical Center of Kosovo, as seen on figure 20, 100% of cases were resistant to the combination of drugs Amoxicillin + Clavulanic Acid, Ampicillin, Norfloxacin, Imipenem and Ciprofloxacin. It is worrisome that in 83.33% of cases (or in 5 cases, out of 6 in total) resistance was encountered towards Vancomycin and Teicoplanin, which are usually stored as antibiotics of last resort.

**Figure 20:**
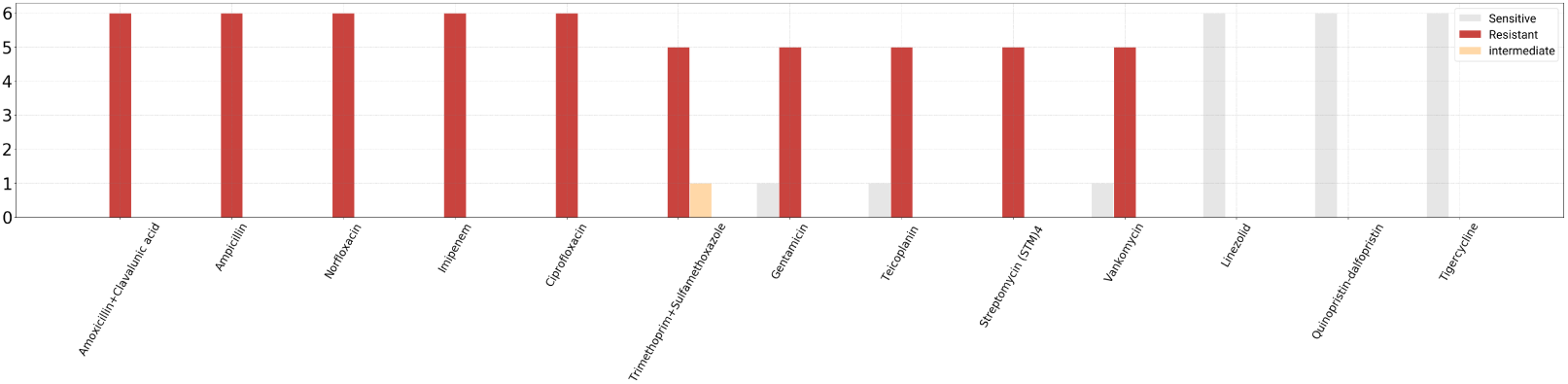
Levels of resistance and sensitivity of Enterococcus faecium to relevant agents

Staphylococcus haemolyticus is one of the most common etiological factors of staphylococcal infections [28]. It has been estimated to be highly resistant, and the reason for this is thought to be the high plasticity genome of the bacterium [28]. Also in our 2 isolated cases,as shown on figure 21, this pathogen has been found to be resistant to almost all antibiotics tested. In 50% of cases it was found to be resistant to: TMP + SMX and Amoxicillin + Clavulanic Acid, Gentamicin, Fusidic Acid, Levofloxacin, Clindamycin, Erythromycin, Moxifloxacin and Norfloxacin combinations. While in 100% of cases, resistance was encountered in: Penicillin, Nitrofurantoin, Aztreonam, Oxacillin and Cefoxitin.

**Figure 21:**
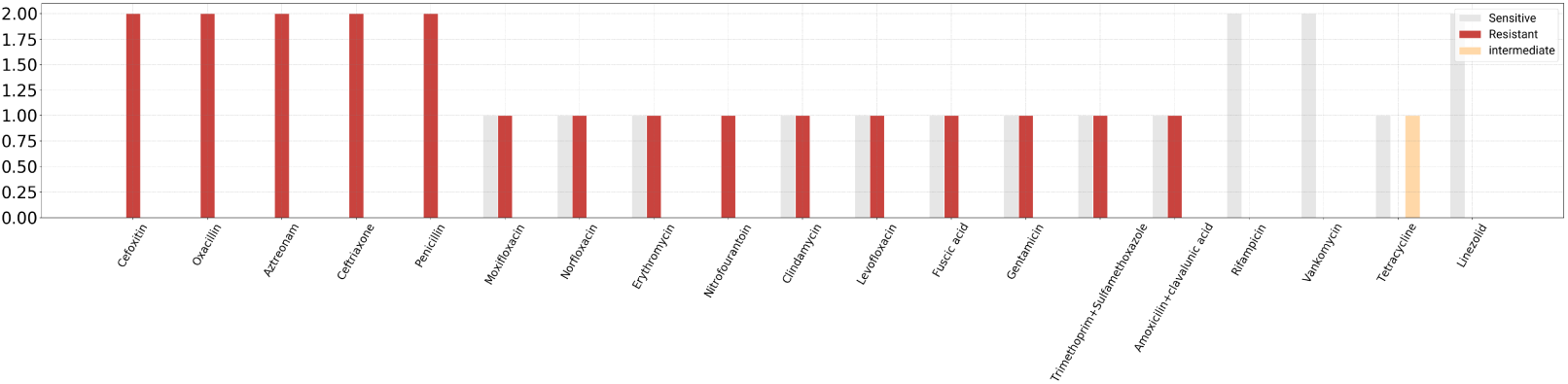
Staph.Haemolyticus resistance and sensitivity levels to relevant agents

## 5. Discussion

In this research, a total of 1499 samples were taken over a 5-year period, of which from only 185 microorganisms (bacteria) were isolated. The cases are mainly from: the infectious disease clinic, the neurology clinic and the neonatology ward from the pediatric clinic. As the research is mainly based on the determination of the etiological profile and the statistics of the occurrence of resistance to the respective antimicrobials, the data does not include information about patients in the above mentioned clinics.

From the results of this research (2014-2019), it has been noticed that the most often isolated bacteria from microbiological samples are: Acetinobacter Baumannii and Staphylococcus spp. Whereas in 2014, according to the annual report of CAESAR (Central Asian and Eastern European Surveillance of Antimicrobial Resistance), in the microbiological samples of CSF, the most commonly isolated bacteria in Kosovo were: Streptococcus Pneumoniae and Pseudomonas aeruginosa. Also according to the same report, it is considered that most samples were taken from the neonatology ward of the pediatric clinic. While in the annual report of CAESAR for 2017 (seen as that for 2018/19, the report will be drafted next year), the most common bacteria isolated from CSF in Kosovo were: Streptococcus Pneumoniae and Enterococcus Faecium. Also this year, most samples of bacterial isolates from CSF were taken from the neonatology department of the pediatric clinic.

- **E.COLI-** from isolated cases with Escherichia Coli, high resistance was encountered in Aminoglycosides (62.5% in Gentamicin, 62.5% in Tobramycin), as well as in third generation Cephalosporins (62.5% in Ceftriaxone, 62.5% in Ceftazidime and 62.5% in Cefotaxime).E.Coli strains in Kosovo (in 2014 according to the annual report of CAESAR-WHO 2014) were 39% resistant to Aminoglycosides, and 48% resistant to third generation of Cephalosporins. While according to CAESAR-WHO 2017, in Kosovo, the resistance of bacteria to Aminoglycosides had increased to 47%, while towards third-generation Cephalosporins was 47%.While in the countries in the region for 2014 (in Serbia), the resistance of E.Col to Aminoglycosides was 33%, while for the third generation Cephalosporins it was also 33%. Meanwhile in North Macedonia for the year 2014: resistance to Aminoglycosides was 64%, while in third-generation Cephalosporins was 73%. For the year 2017 meanwhile, in Serbia, E.Coli resistance to Aminoglycosides was estimated to be 1%, while to third generation Cephalosporins was 29%. This year in North Macedonia the resistance was about 0% in Aminoglycosides and 73% in third generation Cephalosporins. In Montenegro, no resistance was found to Aminoglycosides and 70% resistance to third generation Cephalosporins.
- **K.PNEUMONIAE -** showed resistance mainly to third generation Cephalosporins (Cefotaxime with 83.3%, Ceftriaxone 81.82%, Ceftazidime 50%). In 2014, resistance to third-generation cephalosporins, according to CAESAR-WHO was 91%, while in 2017 this number had increased to 97%.In Serbia, in the annual report of CAESAR-WHO for 2014 the resistance of K.pneumoniae to third generation Cephalosporins was estimated to be 89%. In North Macedonia, this number was lower - 82%. Whereas for the year 2017 the numbers were: Serbia - 84%, North Macedonia - 81%, Montenegro - 97%.
- **ACETINOBACTER SPP -** Most of the isolated cases in this period of 5 years were with Acetinobacter spp (most: Acetinobacter baumannii). Acetinobacter spp being nosocomial pathogens, demonstrate high numbers of resistance. For the last 5 years, the highest resistance has been encountered in the class of Aminoglycosides (Gentamicin with 100%, Amikacin with 100%, Tobramycin with 46%) of antibiotics and in Carbapenems (Imipenem with 93.33%, Meropenem with 100%). In 2014, resistance to aminoglycosides was 96%, while to carbapenems was 90%. In 2017, resistance to aminoglycosides was 93%, while to Carbapenems, it was 89%. According to CAESAR-WHO 2014, in Serbia, the resistance of Acetinobacter spp isolated from CSF samples to Aminoglycosides reaches the number of 93%, and also to Carbapenems 93%. In North Macedonia, this year the resistance to Aminoglycosides is 88%, while to Carbapenems is 65%. In 2017 in Serbia the resistance was: towards Aminoglycosides reached 94%, and towards Carbapenems 95%. In North Macedonia, in the same year the resistance to Amino-glycosides was 82%, and to Carbapenems was 86%. In Montenegro, in 2017 according to CAESAR-WHO, resistance to Aminoglycosides and Carbapenems was 90%.
- **P.AERUGINOSA -** In a total of 12 isolated cases of Pseudomonas aeruginosa, we encounter marked resistance to the antibiotics Carbapenems (Imipenem with 83.33% and Meropenem with 83.33%), as well as to the antibiotics Cephalosporins (second and third generation). In 2014, P. aeruginosa’s resistance in Kosovo to Carbapanems was estimated to be 64%, and to Cephalosporins was 50%. In 2017, resistance to Carbapenems became 74%, while to Cephalosporins it was around 32-42%.According to CAESAR-WHO 2014, in Serbia the resistance of CSF samples towards Carbapenems was 45%, while towards Cephalosporins it was 49%. In North Macedonia, the resistance towards Carbapenems was 38%, while towards Cephalosporins it was 100%. According to the annual report for 2017, in the countries in the region, the resistance statistics were: Serbia-resistance to Carbapenems was 52%; to Cephalosporins 55%, Montenegro-resistance to Carbapenems 36%; to Cephalosporins 38-55%, North Macedonia - 29% to Carbapenems; 24-33% to Cephalosporins.
- **S.AUREUS -** this has been shown to be highly resistant to the macrolide group of antibiotics (100% resistance to Clarithromycin, Azithromycin and Erythromycin). Also, its high resistance is encountered in the antibiotic Penicillin (100% of cases). The percentage of MRSA during this research period was 50% (according to WHO standards, MRSA is calculated as resistance towards the antibiotic cefoxitin). In 2014, according to CAESAR-WHO, the percentage of MRSA in Kosovo was 38%, while in 2017 this number was 58%. In the countries in the region (according to WHO-CAESAR 2014) MRSA has been: in Serbia 33%, in North Macedonia 37%. Whereas in 2017 it was: 26% in Serbia, 23% in Montenegro, and 53% in North Macedonia.
- **S.PNEUMONIAE -** It is worth noting that the isolated S.Pneumoniae bacteria, have shown resistance to a few drugs (multidrug resistance-MDR). The greatest resistance was encountered in the TMP + SMX combination (50% of cases turned out to be resistant). Also in 33.33% of cases we saw resistance towards the antibiotics Clindamycin, Erythromycin, Cloxacillin and Norfloxacin. In 2014, MDR was 0%, while in 2017 it turned out to be 25%. In the countries in the region (according to WHO-CAESAR 2014) MDR has been: in Serbia 23%, in North Macedonia 0%. Whereas in 2017 it was: 23% in Serbia, 25% in Montenegro, and 83% in North Macedonia.
- **E.FAECIUM -** In 100% of cases we have encountered resistance to the combination of drugs Amoxicillin + Clavulanic Acid, Ampicillin, Norfloxacin, Imipenem and Ciprofloxacin. It is disturbing that in 83.33% of cases (or in 5 cases, out of 6 in total) resistance was encountered in Vancomycin and Teicoplanin, which are usually stored as antibiotics of last resort. In the annual report of 2014, the resistance towards Vancomycin was 50%, while in 2017 it was 25%.In the countries in the region (according to WHO-CAESAR 2014) the resistance towards Vancomycin has been: in Serbia 61%, in North Macedonia 65%. Whereas in 2017 it was: 46% in Serbia, 33% in Montenegro, and 52% in North Macedonia.

These data confront us with the frightening truth: antimicrobial resistance is a worldwide crisis for which we do not have a defined, international and comprehensive remedial plan. The European Union was quick to recognize the imminent crisis, and set out methods to monitor antibiotic use and resistance more than 15 years earlier [17]. The European Union systematically evaluates the use of antibiotics and data on resistance, and achieves this despite the great diversity of resources, languages and policies. It should be noted that the United States does not have such a data system, which can be compared to that of the European Union.

The “antibiotic rescue” chain would include the following steps:

1. Creating a rough database for the use and resistance of antimicrobials in the US.
2. Restricting the use of antibiotics in agriculture.
3. Changing old diagnostic methods in microbiology, and implementing new specific target methods.
4. Prevention of nosocomial infections, reinforcing systematic implementation of plans, based on past cases.
5. Aggressive management of antibiotics.
6. Reducing the barrier of the FDA (Food and Drug Administration).
7. Avoiding unnecessary use of antibiotics for viral infections.
8. Stimulating the development of new antibiotics, by pharmaceutical companies.

## 6. Conclusion

- From the results of this paper, we can conclude that resistance to various antimicrobial agents is a very big and widespread problem in the health of our country, especially in cases and in the neonatal ward.
- Of all samples taken and tested of CSF, in only 12.3% of cases microorganisms were isolated.
- Of all the isolated cases, the most common strains were Acetinobacter baumannii and Staphylococcus spp.
- Acetinobacter baumanii was undoubtedly the most isolated bacterium during this 5-year period fromCSF (44 such cases in total). There are two reasons for this: it is a pathogenic opportunistic bacterium, encountered in the hospital environment (nosocomial infection). The most pronounced resistance was found towards Carbapenems (Imipenem with 97.73%, Ertapenem with 100%, Aztreonam with 100%), Cephalosporins (Cefixime, Cefuroxime, Cefoxitin, Ceftazidime, Ceftriaxone - all with 100%), Aminoglycosides with 100%, Tobramycin with 100%).
- The multidrug resistance of Acetinobacter spp. to various antimicrobial agents has become problematic, and could potentially turn into nonresistance. In our research, Acetinobacter spp., have been shown to be sensitive to Colistin, Minocycline and Tigecycline.
- In 20 cases where Staph. Spp were isolated from CSF samples, the highest resistance (95%) was encountered in Penicillin. Macrolide antibiotics Clarithromycin and Azithromycin with 77.78% were not left behind.
- It is disturbing that in isolated cases of Pseudomonas aeruginosa, a marked resistance towards the Carbapenem antibiotics was encountered (Imipenem with 83.33% and Meropenem with 83.33%), as well as towards Cephalosporins (second and third generation) antibiotics.
- In this period of 5 years the facultative anaerobic bacterium, Listeria Monocytogenes, has not been shown to be resistant to the antibiotics in which it has been tested. In all tests it turned out to be sensitive.
- High levels of resistance of Enterococcus Faecium are alarming. It is especially surprising that in Kosovo this bacterium has already gained resistance in Vancomycin (83.33%), Teicoplanin (83.33%) and Streptomycin (100%), therapies that are usually left for last resort. They are still sensitive to Linezolid and Tigecycline.

## 7. Summary

In CNS infections, the laboratory workup of CSF is considered to be of essential value in order to validate a diagnosis, identify the causative microorganisms and pave the road to antimicrobial therapy. Bacterial infections in the central nervous system are: Bacterial meningitis (the most common one), brain abscess, encephalitis and spinal epidural abscess.

The discovery of antimicrobial agents is considered to be a turning point towards revolution in the history of modern medicine. Yet, inadequate application, inaccurate patient-informing, application of antimicrobial agents outside the medical sector, inability to discover new and more sensitive antimicrobial agents, have directly resulted in the development of bacterial resistance towards various antimicrobial agents. Many bacterial strains have developed multi-resistance to many agents, through various genetic mechanisms of action, contributing to this global problem, through the process of natural selection.

The purpose of this research was to recognise the etiologic profile of CNS infections (especially meningitis), as well as the recognition of the degree of microbial resistance. This research is a retrospective analysis, which contains the information in the time-period from January the first,2014 up to May the 7th, 2019. Overall, there were 1499 samples, from which 185 bacteria were isolated and there were 1314 non-isolates.This data was provided by the National Institute of Public Health in Kosovo. Determination of antimicrobial resistance at the Public Health Institute in Kosovo was carried out by the automated systems and the disk diffusion method.

These study samples are part of the CAESAR study, which is a key ring of the World Health Organization (WHO), which collects data for sensitivity and resistance testing in antibiotics from relevant samples for some of the bacterial species that have a greater clinical relevance.

High resistance levels in 3rd and 4th generation Cephalosporins as well as Aminoglycosides were a common occurrence for all of the isolates in this study. Acinetobacter spp. have been Isolated more frequently, and at the same time, were found to be more resistant to many drugs (MDRs).

High resistance levels in E.faecium, P.Aeruginosa and Acinetobacter spp are quite concerning and may be a reflection on the dissemination of resistant strains in the health care facilities.

## Data Availability

The dataset is also included in the Supplemental Materials. All data for this study can also be obtained via the provided public google drive link.

https://docs.google.com/spreadsheets/d/1NSId2JD2t_HIUGyYxh7EzTw5HkBOvzCbEO9M6chcd7k/edit?usp=sharing

